# Genomically Adjusted Radiation Dose Predicts Outcomes in Pediatric Brain Tumors and Supports Biologically Personalized Radiotherapy

**DOI:** 10.64898/2026.08.09.26359976

**Authors:** Nikhil Joshi, Drew Bergman, Shivani Nellore, Peng Chen, Erin Murphy, Saad Sheikh, Michael LaRiviere, Jessica Foster, Jill Durkin, Anuoluwapo Ajao, Thomas Matulis, Ronica Nanda, Kosj Yamoah, Stacie Stapleton, Chris Beltran, Steven A. Eschrich, Javier F. Torres-Roca, Jacob G. Scott

## Abstract

**Background:** Radiotherapy is a cornerstone of treatment for pediatric central nervous system (CNS) tumors, but dose selection remains uniform despite interpatient variability in tumor radiosensitivity. This is consequential in children, for whom toxicity can have lifelong effects. Genomic-adjusted radiation dose (GARD) integrates tumor genomics with delivered radiation dose to quantify the biological effect of radiotherapy and has been validated across adult malignancies. Its relevance in pediatric CNS tumors is unknown.

**Methods:** We performed a cohort study using gene expression and data from 246 pediatric patients with high-grade glioma, medulloblastoma, or ependymoma from the Children’s Brain Tumor Network. GARD was calculated using a sequencing-adapted radiosensitivity index integrated with radiation dose through the linear–quadratic model. Associations of GARD and physical radiation dose with event-free and overall survival were evaluated using Cox proportional hazards models stratified by tumor type and anatomic location. Patients who did not receive radiotherapy comprised a negative control cohort (sham-GARD).

**Results:** Among patients receiving radiotherapy, physical dose was uniform, whereas GARD showed substantial interpatient variability in biological effect. Higher GARD was associated with improved event-free survival (hazard ratio [HR] 0·90, 95% CI 0·83–0·97; p=0·004) and overall survival (HR 0·90, 95% CI 0·83–0·99; p=0·018). Physical dose was not associated with either endpoint. Among patients who did not receive radiotherapy, sham-GARD was not associated with outcomes, supporting GARD as a treatment-specific predictor rather than a prognostic biomarker.

**Conclusions:** In pediatric CNS tumors, the biological effect of radiotherapy quantified by GARD was associated with outcomes, whereas physical dose was not. These findings challenge uniform radiotherapy dosing and support genomically informed dose individualization. Prospective evaluation of GARD-guided radiotherapy is warranted to optimize tumor control while minimizing long-term toxicity in children.

## Introduction

Pediatric central nervous system (CNS) tumors represent the most common solid tumor malignancy in childhood and remain the leading cause of cancer-related mortality in children. [1, 2] The management of these tumors is uniquely challenging given their frequent proximity to eloquent and developmentally critical brain structures. Consequently, treatment is typically multi-modal incorporating surgical resection, radiation therapy, and/or systemic or targeted therapies to maximize tumor control. [3, 4] While this comprehensive care paradigm has substantially improved survival outcomes, radiation is also associated with significant long-term late effects that can profoundly impact neurologic function, development, and overall quality-of-life throughout survivorship. [5] As such, optimal management of pediatric CNS tumors requires careful balancing of tumor-directed efficacy with preservation of long-term neurologic and functional outcomes.

Radiation therapy remains a cornerstone of treatment for many pediatric CNS tumors but is also a major contributor to treatment-related late effects, including neurocognitive impairment, radiation-induced secondary malignancies, and developmental sequelae that may persist or evolve over decades of survivorship. [5–7] These risks impose significant challenges with the use of radiotherapy in pediatric populations and have driven substantial technological innovation aimed at reducing normal tissue exposure, including advanced conformal techniques and heavy ion-beam radiotherapy. [8] Despite these technical advances in radiation delivery, radiotherapy dosing paradigms have remained largely uniform, with a one-size-fits-all approach still applied in pediatric CNS tumors. [9, 10] Importantly, while radiotherapy is administered in a standardized manner, its biological effect does not follow a similarly uniform pattern, underscoring the need for dosing strategies that account for interpatient heterogeneity in radiotherapy response. [11]

Large-scale studies have suggested that this heterogeneity is driven, in part, by differences in tumor genomics that influence intrinsic radiosensitivity. [12, 13] Therefore, many efforts have sought to develop surrogate measures of individual tumor radiosensitivity to predict therapeutic response. [11, 14–17] In pediatric oncology, however, prior work has largely focused on classifying into clinical or histological subgroups that may benefit from radiotherapy, rather than quantifying patient-specific biological response. [18–20] To date, no studies to our knowledge have comprehensively characterized the relationship between tumor genomics, predicted intrinsic radiosensitivity, and radiation response in pediatric CNS tumors.

To address these issues more broadly, our group previously developed the gene expression–based radiosensitivity index (RSI), a robust 10-gene classifier that stratifies tumors according to intrinsic radiosensitivity. [12, 21] RSI has been validated across multiple independent cohorts and diverse cancer types. [13, 16, 22, 23] Building on this work, we developed the genomic-adjusted radiation dose (GARD), which integrates RSI with physical radiation dose to quantify a predicted biological effect of radiotherapy in an individual patient. [24] GARD has likewise been validated across numerous adult cancer cohorts and provides a biologically grounded framework for estimating the expected therapeutic benefit of a given physical dose based on tumor-intrinsic radiosensitivity. [11, 14, 15, 25, 26] Importantly, GARD also offers a prescriptive approach to radiation delivery, allowing clinicians to tailor the physical dose to achieve a desired biological effect in a patient-specific genomically informed manner. Together, these efforts establish a direct link between tumor genomics, dose and outcome.

Building on extensive validation in adult malignancies, we sought to characterize the relevance and clinical utility of GARD-based radiotherapy dosing paradigms in pediatric CNS tumors. Utilizing gene expression and clinical data from the Children’s Brain Tumor Network (CBTN), we evaluated pediatric high-grade gliomas, medulloblastomas, and ependymomas to examine the relationship between radiation-intrinsic biological effect, as quantified by GARD, and clinical outcomes including overall survival and time to first recurrence. Through this work, our aim is to provide a foundation for genomically-informed radiotherapy personalization in pediatric neuro-oncology.

## Methods

### Study Design

Data were collected from samples available through the Children’s Brain Tumor Network’s (CBTN) biorepository. [27] Samples with sufficient information (gene expression, radiation timing, dose and fractionation, and survival metrics) to calculate GARD and clinical outcome were included. In cases where dose per fraction was unavailable, this value was imputed using standard disease-specific schedules reported in cooperative-group studies of pediatric medulloblastoma, high-grade glioma, and ependymoma. [10, 28–30] Samples selected for analysis fell into three distinct tumor categories: high-grade glioma, ependymoma, and medulloblastoma. These three tumor types were comprised of 246 total patients at initial diagnosis with 207 (84.1%) definitively treated with radiotherapy and 39 (15.9%) not treated with radiotherapy. Untreated patients were included as a negative control to determine if GARD is specific only among patients treated with radiotherapy. Clinical and demographic information for the cohort is described in **Supplemental Table 1**. Clinical endpoints included time to first recurrence (EFS) and overall survival (OS), each defined as the interval from the date of pathological diagnosis to the respective outcome.

### GARD Calculation

Genomic Adjusted Radiation Dose (GARD) was calculated as described previously, using the linear–quadratic (LQ) model in combination with each patient’s radiation dose and fractionation schedule, as well as their individualized radiosensitivity index (RSI). [24] The RSI model was originally developed using gene expression data from 48 cancer cell lines and was trained to predict intrinsic radiosensitivity as quantified by the survival fraction following 2 Gy of radiation (SF2). The original RSI algorithm incorporates ten genes; each ranked according to relative expression within a sample (with ranks assigned from 1 for lowest expression to 10 for highest expression). [12]

Given that this model was developed using Affymetrix gene arrays and thus only applies to similar platforms, we established a transformation of the original RSI model that may be applied to traditional short-read RNA-sequencing gene expression values (hereafter referred to as RSI*_seq_*). [31] RSI*_seq_*identified the appropriate numeric radiosensitivity in each tumor using the following formula:

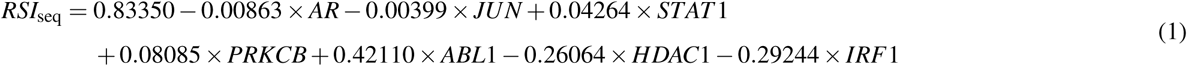

A patient-specific radiosensitivity was estimated by computing RSI*_seq_* by substituting normalized transcripts-per-million quantities in the equation above. To compute GARD, RSI*_seq_*was substituted for the survival term in the standard linear quadratic equation. This yielded the following:

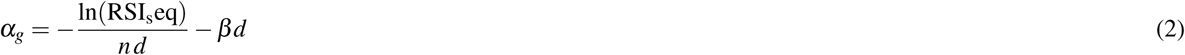

where the dose per fraction (*d*) is set to 1.8 Gy, the number of fractions (*n*) is equal to 1, and *β* is assumed to be 0.05/Gy^2^. These values align with GARD computation procedures from previous works. Finally, GARD is calculated by applying the equation for biological effect with patient-specific *α_g_* calculated as per equation (3). All other constants remained the same.

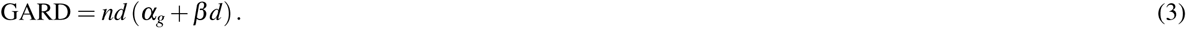

### Statistical Analyses

All analyses were completed in R 4.4.2. GARD calculation was computed as previously reported. To ensure biologically appropriate comparisons, all doses are converted to EQD2 using an 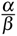 of 10; however, these values are referred to as physical or total radiation dose for ease of interpretation. Given that sequencing data was abstracted from bulk-RNA-sequencing data, spatial intratumoral heterogeneity was not considered. Future analyses will interrogate this important correlate.

Analyses relating GARD to outcome measures were completed using Cox proportional hazards models. To complete pooled analyses across distinct tumor types and brain regions, we utilized a Cox proportional hazards model stratified by brain region and cancer type with GARD, physical radiation dose or sham-GARD as the only covariates. Utilizing this method allowed us to compute baseline GARD hazard models while also allowing us to compute a pooled GARD hazard model considerate of all pediatric brain tumors together. Cox proportional hazards assumptions were tested and 5-year overall survival and event-free survival were selected as endpoints to ensure hazards were invariable over time. Proportional hazards assumptions were evaluated using Schoenfeld residuals.

Kaplan-Meier survival analysis was utilized to further relate GARD to outcome measures. Cut point analysis for GARD was performed using maximally selected rank statistics with the *surv*_*cut point*() function (survminer package, R), which identifies the threshold that maximizes separation of survival outcomes. This was completed on a per-tumor basis. Dichotomized Cox-proportional hazards models applied these cutpoints to determine GARD categories.

To ensure patients included in analysis were being treated with curative intent and not receiving palliative radiotherapy, we selected samples that fell within the standard of care dose range for dose administration (50 to 60 Gy administered). This range encompasses commonly used focal or boost doses in the disease-defining pediatric trials included in this analysis. [28–30] In cases where the total dose was unknown and definitive radiotherapy was received, we assumed patients received the standard-of-care radiation dose (54 Gy in 30 fractions). We acknowledge this is a strong assumption, however, we note that this dose fell within the interquartile range of observed doses and aligned closest with standard-of-care regimens supporting its biological and clinical plausibility.

The pooled analysis was completed by utilizing a stratified Cox proportional hazards model with stratification by cancer type and brain region and, separately, a *χ*-square statistic calculated utilizing the Wald test. Patients who did not receive radiotherapy were pooled into a cohort called sham-GARD for a similar analysis. We modeled sham-GARD by assigning the standard-of-care dose of RT to this cohort (54 Gy in 30 fractions) and computing GARD as previously described.

## Results

### Intrinsic Radiosensitivity Reveals Significant Histologic and Molecular Heterogeneity Across Pediatric CNS Tumors

To understand radiosensitivity of pediatric CNS malignancies, we initially determined the intrinsic radiosensitivity (RSI) for each patient receiving radiotherapy. Of note, tumor radiosensitivities aligned closely with previously described tumor-intrinsic radiosensitivity distributions. We noted a heterogenous distribution of radiosensitivities across histologies with marked differences between medulloblastoma, high-grade glioma, and ependymal tumors (Kruskal-Wallis p < 0.001) (**Supplemental Figure 1A**). Molecular subgroups revealed limited subclass-specific differences in radiosensitivity (**Supplemental Figure 2**). Furthermore, brain region-based stratification identified significant differences in radiosensitivity independent of tumor histology (**Supplemental Figure 1B**). Even after accounting for tumor histology and brain region, however, substantial inter-sample heterogeneity in radiosensitivity remains (**Supplemental Figure 1C**). We therefore applied GARD to interrogate tumor radiosensitivity with delivered dose and assess whether this approach contextualizes heterogeneity in predicted radiotherapy effect across pediatric brain tumors.

### GARD Reveals Inter-Patient Variability in Predicted Radiotherapy Effect

GARD values were calculated for each patient receiving radiotherapy. Physical radiation dosing aligned closely to standard of care regimens with most patients receiving 54 Gy in 30 fractions or 59.4 Gy in 33 fractions. While limited variability in physical radiation dose persisted, there was a varied distribution of expected biological effects across tumor types – highlighting disparate biological outcomes under similar dosing paradigms (**Figure 1, Supplemental Figure 3**). GARD distributions were not significantly associated with patient age or sex (**Supplemental Figure 4**). Among patients with medulloblastoma, GARD distributions also did not differ between conventional high-risk and low-risk treatment groups, which receive craniospinal irradiation doses of approximately 36 Gy and 23.4 Gy, respectively (p = 0.87).

**Figure 1.**
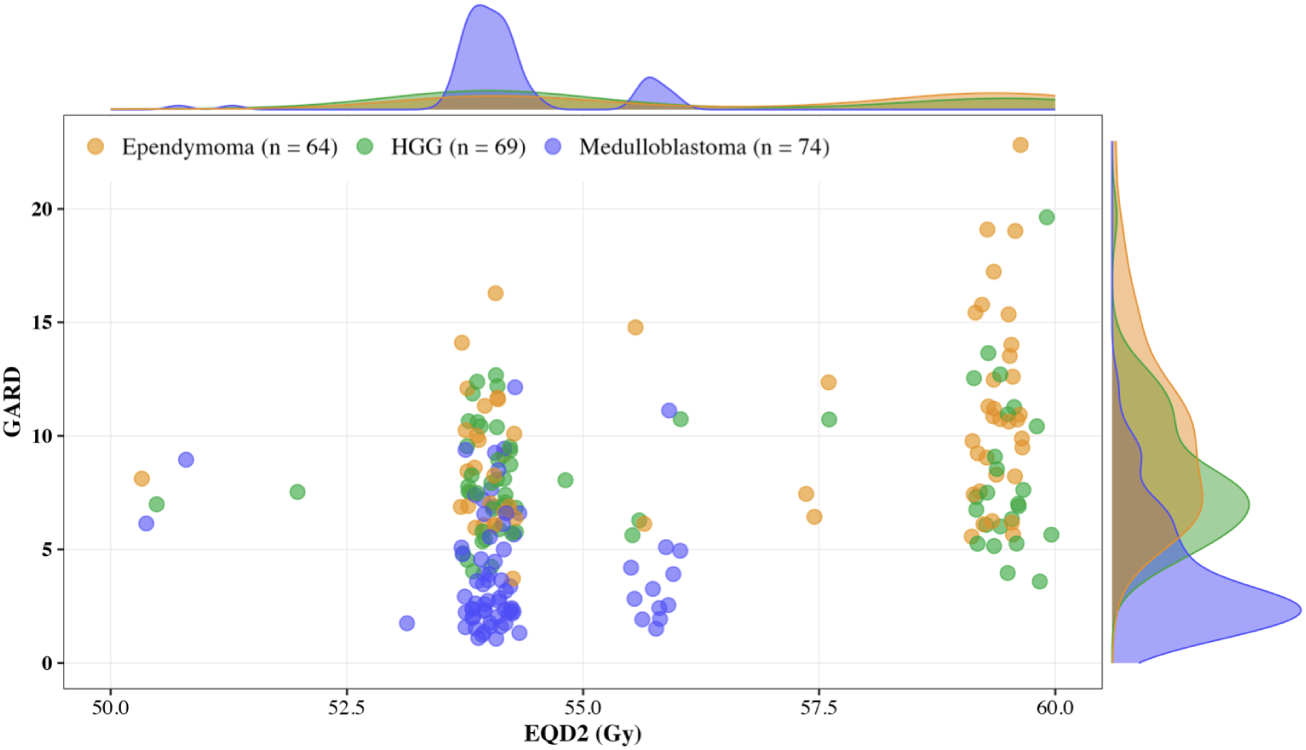
Patient-level association scatter plot of physical dose (EQD2) of radiation delivered versus GARD. While EQD2 for each disease site is relatively uniform, GARD presents wide variability. All cancer types are included and colored by histology. Samples are filtered to between 50 and 80 Gy. Kernel densities for each variable are present on the margins.

### GARD Is Associated with Survival Outcomes in Patients Receiving Radiotherapy

To evaluate the clinical relevance of GARD, pooled survival analyses were performed in patients who received radiotherapy. Pooled analyses using Cox proportional hazards demonstrated that GARD is associated with time to first recurrence (HR = 0.90 [0.83-0.97], **p = 0.004**) and overall survival (HR = 0.90 [0.83-0.99], **p = 0.018**) in patients who received radiotherapy (**Figure 2 A, 2C**). This relationship was preserved when including histology and brain region as covariates within the model (**Supplemental Figure 10**, **Supplemental Table 5**). Dichotomized Cox-proportional hazards modeling stratified by histology and brain region demonstrated similarly improved outcomes for patients with high-GARD against low-GARD patients for both event-free (HR = 0.56 [0.37-0.84], **p = 0.005**) and overall (HR = 0.56 [0.35-0.88], **p = 0.013**) survival (**Figure 3**).

**Figure 2.**
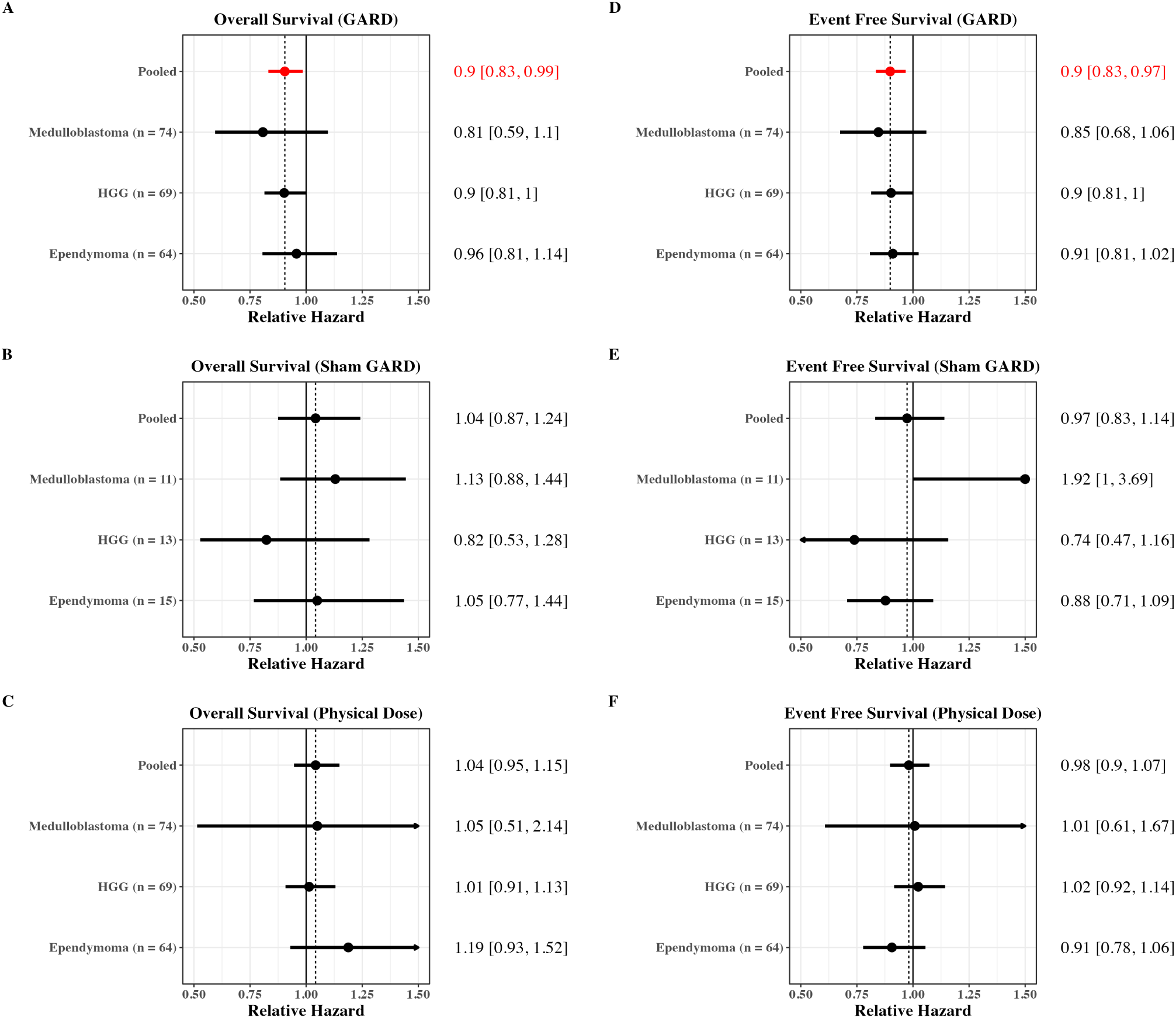
Individual Cox proportional relative hazards for each cancer type across event-free and overall survival. **(A)** Cox proportional hazards models with GARD for overall survival for each tumor type separately and for the pooled analysis across tumor types. **(B)** Cox proportional hazards models with sham-GARD for overall survival for each tumor type separately and for the pooled analysis across tumor types. **(C)** Cox proportional hazards models with physical dose of radiation as covariate for overall survival for each tumor type separately and for the pooled analysis across tumor types. **(D)** Cox proportional hazards models with GARD as covariate for event-free survival for each tumor type separately and for the pooled analysis across tumor types. **(E)** Cox proportional hazards models with sham-GARD as covariate for event-free survival for each tumor type separately and for the pooled analysis across tumor types. **(F)** Cox proportional hazards models with physical dose of radiation as covariate for event-free survival for each tumor type separately and for the pooled analysis across tumor types. Data for relative hazards and 95% confidence intervals are rounded to two decimal places for ease of presentation. Dashed vertical line delineates the pooled relative hazard. Red coloring indicates significant findings. GARD = Genomic-adjusted radiation dose.

**Figure 3.**
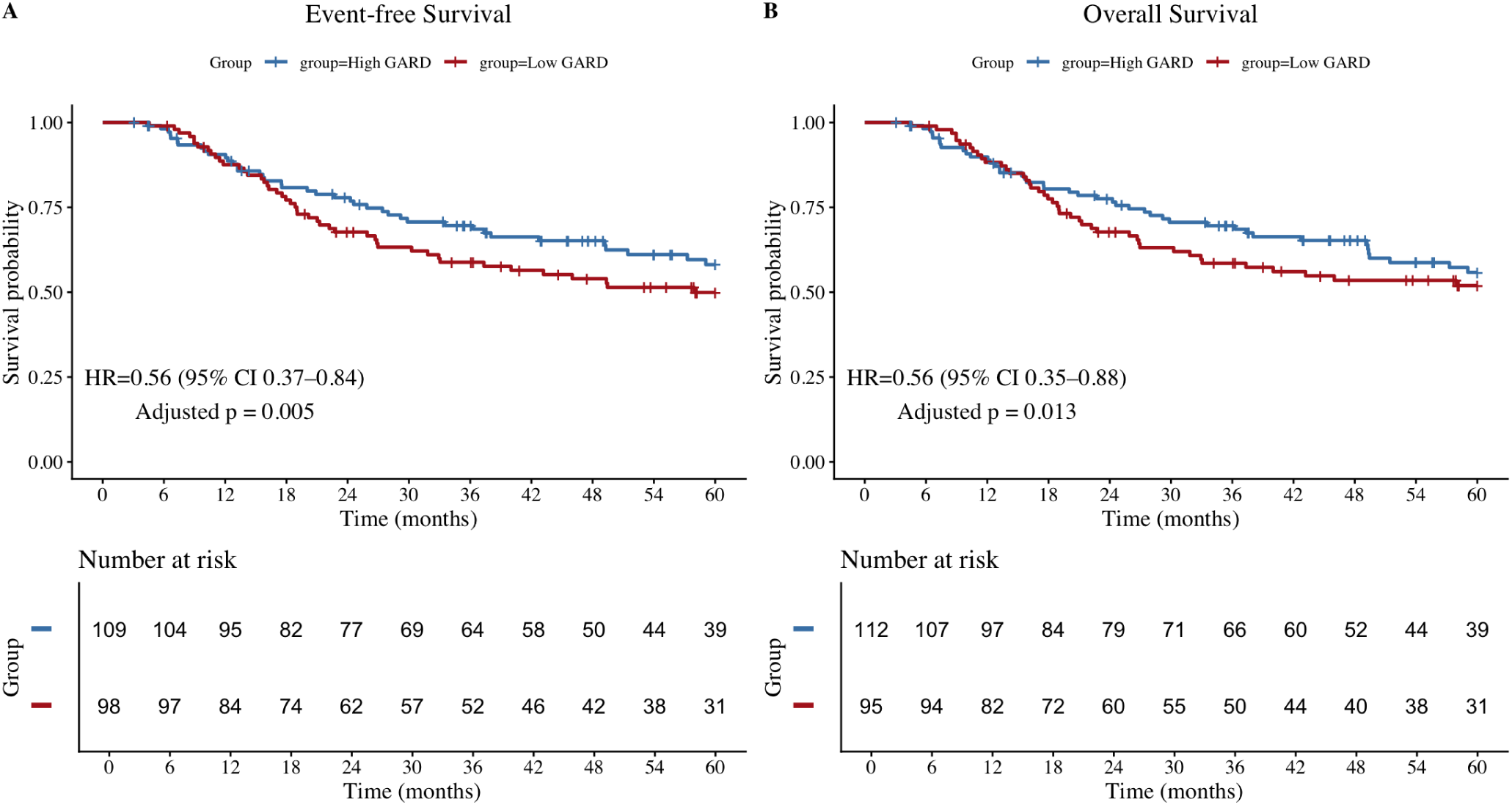
Event-Free and Overall Survival Stratified by Dichotomized GARD. **(A)** Kaplan–Meier curve for event-free survival. **(B)** Kaplan–Meier curve for overall survival. Hazard ratios and associated statistics were calculated using Cox proportional hazards models comparing GARD-low with GARD-high patients and stratified by histology and brain region.

To assess whether GARD functioned as a general prognostic biomarker or a treatment-specific predictor, parallel analyses were performed in control cohorts of patients who did not receive radiotherapy (sham-GARD). In contrast to radiotherapy-treated patients, GARD demonstrated no association with time to first recurrence (HR = 0.97 [0.83, 1.14], p = 0.737) or overall survival (HR = 1.04 [0.87, 1.24], p = 0.638) in patients not treated with radiotherapy (**Figure 2B**, **Figure 2E**). There was no association found between physical dose of radiotherapy and time to first recurrence (HR = 0.98 [0.90, 1.07], p = 0.676) or overall survival (HR = 1.04 [0.95, 1.15], p = 0.395) (**Figure 2C, 2F**, **Supplemental Table 3**).

GARD demonstrated a significant linear association with both time to recurrence and overall survival (**Figure 4**). Increasing GARD values were associated with progressively lower GARD-specific relative hazards for both recurrence and mortality (time to first recurrence: p = 0.0037; overall survival: p = 0.0185). Evaluation of clinical outcomes at the 5-year timepoint demonstrated that GARD functioned as a continuous predictor of both recurrence risk and survival probability. Specifically, increasing GARD values were associated with reduced 5-year recurrence probability and improved 5-year overall survival among radiotherapy-treated patients.

**Figure 4.**
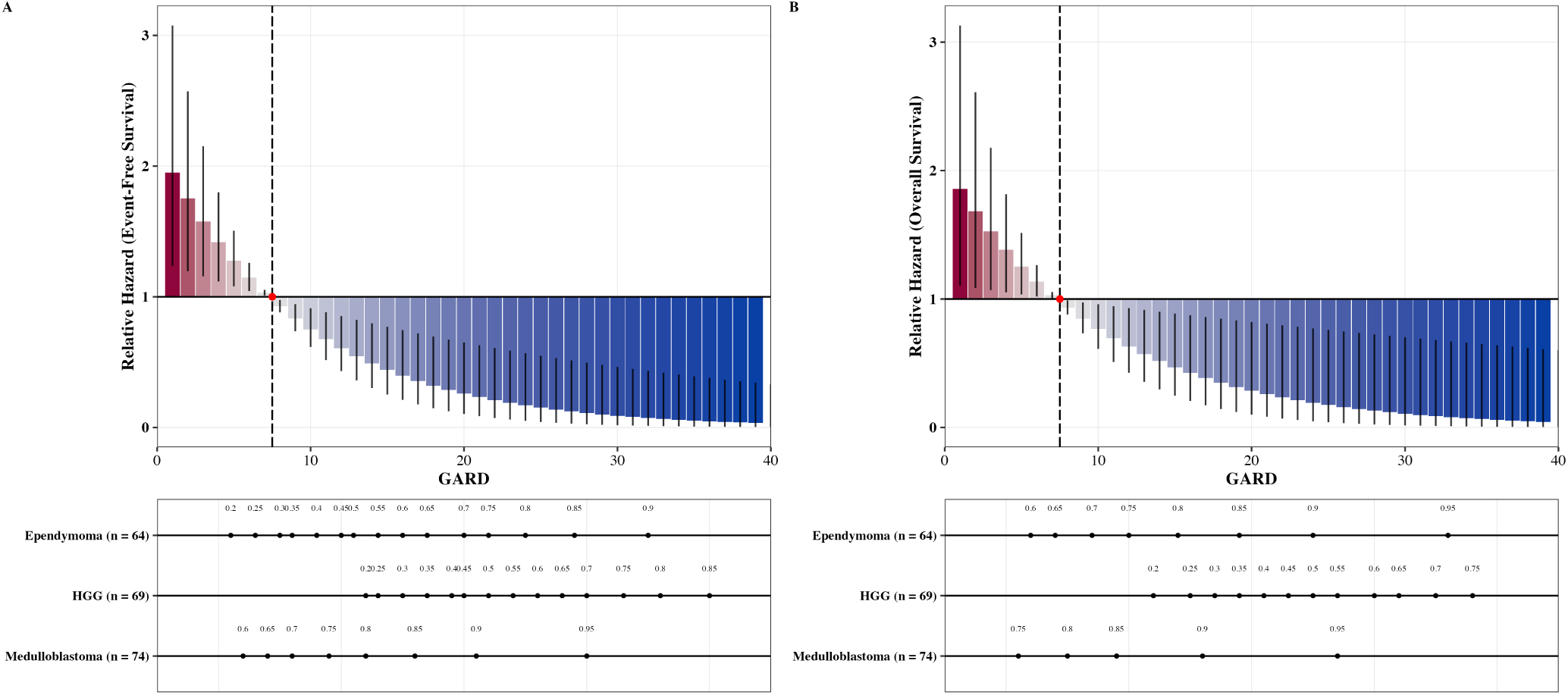
Relative hazard per unit GARD at 5 years after diagnosis as predicted by the stratified Cox model for both (A) event-free survival and (B) overall survival by cancer site and brain region. Nomograms (bottom) indicates the effect of relative hazard as determined by GARD in the context of a specific cancer type and the predicted absolute survival probability underlying each corresponding GARD value. The red dot and vertical line indicate the mean GARD for the cohort. The red to blue gradient depicts the gradient from highest (red) to lowest (blue) hazard for this continuous analysis. Error bars are 95% confidence intervals. GARD = genomic-adjusted radiation dose.

## Discussion

Radiotherapy remains a cornerstone of treatment across many pediatric brain tumors, yet current dosing frameworks largely rely on uniform, one-size-fits-all paradigms. [10] In this pooled analysis, we found that the biological effect of radiotherapy was associated with longer time to recurrence and improved overall survival among children receiving radiotherapy, whereas prescribed physical dose alone was not. Each 1-unit increase in GARD was associated with a 10% reduction in hazard (HR 0.90), corresponding to an estimated 50% relative reduction in hazard across the interquartile range of observed GARD values. Importantly, these findings suggest that current radiotherapy dosing paradigms effectively distribute patients across a wide range of biological effect despite delivering near-uniform physical doses. In this framework, current dosing paradigms may unintentionally distribute patients across a broad range of biological effects, with some potentially receiving insufficient biological dose and others receiving additional exposure with limited incremental benefit. GARD provides a means to resolve this mismatch by aligning prescribed dose with expected biological effect.

These findings align with prior work showing that the biological effect of radiotherapy, rather than prescribed dose alone, better captures treatment response across diverse malignancies. [15] Pan-cancer analyses have demonstrated that genomically informed radiation-effect metrics (GARD) can predict survival outcomes and distinguish which patients benefit from radiotherapy, emphasizing the role of intrinsic radiosensitivity in treatment efficacy. Our analysis extends this framework specifically to pediatric brain tumors, suggesting that integrating genomic measures of radiation response could improve risk stratification beyond traditional clinical factors such as histology, tumor site, extent of resection, and metastatic status. Clinically, these observations support the potential for biologically guided dosing strategies. Such approaches could permit moderate escalation for predicted radioresistant tumors while enabling safe de-escalation for highly radiosensitive disease, improving tumor control without proportionally increasing toxicity to eloquent structures. Indeed, these biomarkers supply informative decision-support metrics to help determine which patients are likely to benefit from radiation and the level of radiation that should be administered. This balance is especially consequential in children, where treatment decisions have impacts on long-term neurodevelopmental outcomes. [5, 8, 32]

With this context in mind, several limitations warrant consideration when interpreting these findings. Inclusion of multiple histologies limits direct clinical comparability, and current GARD modeling primarily reflects tumor-directed dosing near standard fractionation, restricting assessment of hypofractionation and craniospinal irradiation. Limited statistical power also precluded robust molecular subtype–specific analyses, which remain an important direction for future study. These constraints highlight the need for prospective validation and more granular biologic characterization in future cohorts. Even with these limitations, the results point toward an important opportunity for advancing treatment personalization. By quantifying the expected biological effect of a given dose in an individual tumor, GARD provides a framework for moving beyond conventional histology-driven dosing paradigms towards more individualized radiotherapy planning.

GARD should not be interpreted as a conventional prognostic or outcome-prediction model, as it was neither designed nor trained to directly forecast clinical endpoints. Rather, GARD quantifies the biological effect of a given radiation dose within an individual tumor, integrating intrinsic radiosensitivity with prescribed dose to estimate radiation-induced biological effect. Any observed association between GARD and survival or recurrence therefore reflects its ability to capture inter-patient variation in radiotherapy-specific treatment benefit, not disease prognosis. Its predictive scope is confined to the component of outcome attributable to radiotherapy itself. Although this narrower focus may limit its performance as a global prognostic model, it enhances its clinical relevance: GARD offers a mechanistic estimate of how much radiotherapy is expected to contribute to disease control for a particular patient.

Recent pediatric CNS trials illustrate the increasing integration of tumor biology into radiation strategies. Cooperative-group protocols such as ACNS0331 and SJMB12 demonstrate that molecular and clinical stratification can refine treatment intensity, particularly through risk-adapted craniospinal irradiation (CSI) dosing and subgroup-specific therapeutic arms. [33, 34] More recently, de-escalation trials such as ACNS1422 have investigated reduced CSI dosing in biologically favorable WNT-driven medulloblastoma, reflecting confidence in improved outcomes within this subgroup. [35] These efforts represent a major advance over purely anatomy-based paradigms. However, even within molecularly-defined groups, radiation dose remains prescribed in a uniform manner. Patients within the same subgroup receive uniform focal (e.g. 54 Gy) and CSI doses (e.g. 23.4 Gy or 36 Gy), despite potentially substantial heterogeneity in intrinsic radiosensitivity. Notably, our findings suggest that anatomic location and molecular subtype alone do not fully determine tumor radiosensitivity, with substantial variability persisting even among tumors sharing similar clinicomolecular features. Our findings suggest that this within-subgroup variability may be clinically meaningful.

The observed distribution of radiosensitivity indices and corresponding GARD values within established risk and molecular strata indicate that molecular classification alone does not fully capture differences in radiation response. As pediatric trials increasingly explore molecularly guided dose de-escalation – particularly in favorable biology cohorts – it would be prudent to incorporate biologically informed metrics such as GARD as a pre-treatment decision-support or stratification tool.

A pertinent example of this is applied in risk stratification of medulloblastoma. Given that the biological effect inferred from GARD supposes no difference in low/high-risk groups, there may be patients who are not being optimally stratified into these clinically-relevant treatment axes. In fact, our exploratory analyses suggest that there may be low-risk patients who benefit from increased irradiation and high-risk patients who benefit from reduced irradiation based on inferences from their GARD profile. In this setting and others, using these biomarkers may offer a prudent decision-support tool to determine the optimal radiation dosing paradigm informed by the inherent radiosensitivity of a given patient. In this framework, GARD would not replace existing risk stratification, but rather complement it by identifying patients whose tumors are predicted to derive robust radiation benefit at lower doses, as well as those who may require maintained or intensified dosing despite favorable molecular features. Such integration could allow future de-escalation strategies to move beyond subgroup-level assumptions toward truly individualized radiotherapy personalization.

As a genomic framework designed to estimate radiotherapy benefit, GARD has the potential to play an important role in advancing precision radiation oncology for pediatric brain tumors. Unlike traditional approaches that rely primarily on prescribed physical dose, GARD provides patient- and tumor-specific information that may help clinicians anticipate the magnitude of radiotherapy benefit for an individual tumor. As translational efforts and prospective studies continue to emerge, incorporating biological response metrics such as GARD into pediatric neuro-oncology workflows could help move the field toward more individualized radiotherapy strategies that balance tumor control with long-term survivorship outcomes.

Pediatric radiation oncology has spent the last two decades personalizing where radiation is delivered, how much tissue receives radiation, and which patients require radiation. Underlying this, however, the prescribed tumor dose itself remains largely uniform across disease populations. Our findings suggest that the next evolution in pediatric radiotherapy may come not from delivering radiation more precisely, but from delivering a more appropriate biological effect to each child.

## Data Availability

All data produced in the present study are available upon reasonable request to the authors.

## Supplemental Information

### Code and data availability

Statistical analyses were conducted using R v4.4.2 and associated packages. All analyses R scripts are available at: https://github.com/nyquiliousjoshi/GARD_pediatrics. Open data access is available via the CBTN.

### Cohort clinical characteristics

Given this is a large cohort of patients across age ranges, we describe the clinical features of the patient population described on a per-tumor basis, demonstrating that this cohort is representative of our pediatric brain tumor populations (see **Supplemental Table 1**).

**Supplemental Table 1.** Clinical and demographic characteristics of patients across tumor types. Values are presented as mean (SD) or n (%). ^1^Mean (SD); n (%) ^2^Kruskal-Wallis rank sum test; Pearson’s Chi-squared test; NA

| Characteristic | Ependymoma (N = 79)<sup>1</sup> | HGG (N = 82)<sup>1</sup> | Medulloblastoma (N = 85)<sup>1</sup> | p-value<sup>2</sup> |
| --- | --- | --- | --- | --- |
| Age (years) | 7.1 (5.6) | 10.0 (4.8) | 9.6 (4.8) | < 0.001 |
| **Sex** |  |  |  | 0.5 |
| Female | 34 (44%) | 42 (51%) | 36 (42%) |  |
| Male | 43 (56%) | 40 (49%) | 49 (58%) |  |
| Unknown | 2 | 0 | 0 |  |
| **Radiotherapy** | 64 (81%) | 69 (84%) | 74 (87%) | 0.6 |
| Photons | 7 (8.9%) | 27 (33%) | 6 (7.1%) |  |
| Protons | 41 (52%) | 20 (24%) | 46 (54%) |  |
| Unknown | 16 (20%) | 22 (27%) | 22 (26%) |  |
| **Brain Region** |  |  |  |  |
| Hemispheric | 28 (35%) | 33 (40%) | 0 (0%) |  |
| Midline | 2 (2.5%) | 25 (30%) | 0 (0%) |  |
| Mixed | 13 (16%) | 18 (22%) | 17 (20%) |  |
| Other | 1 (1.3%) | 1 (1.2%) | 1 (1.2%) |  |
| Posterior fossa | 14 (18%) | 3 (3.7%) | 62 (73%) |  |
| Spine | 4 (5.1%) | 1 (1.2%) | 0 (0%) |  |
| Suprasellar | 0 (0%) | 1 (1.2%) | 0 (0%) |  |
| Ventricles | 17 (22%) | 0 (0%) | 5 (5.9%) |  |

### RSI distributions

The distribution of RSI was compared against cancer type and brain region as these features are both known clinically-relevant features with radiotherapeutic dosing relevance. We additionally utilized linear regression modeling to determine the portion of variability in RSI informed by these two critical features. These analyses are included in **Supplemental Figure 1**.

**Supplemental Figure 1.**
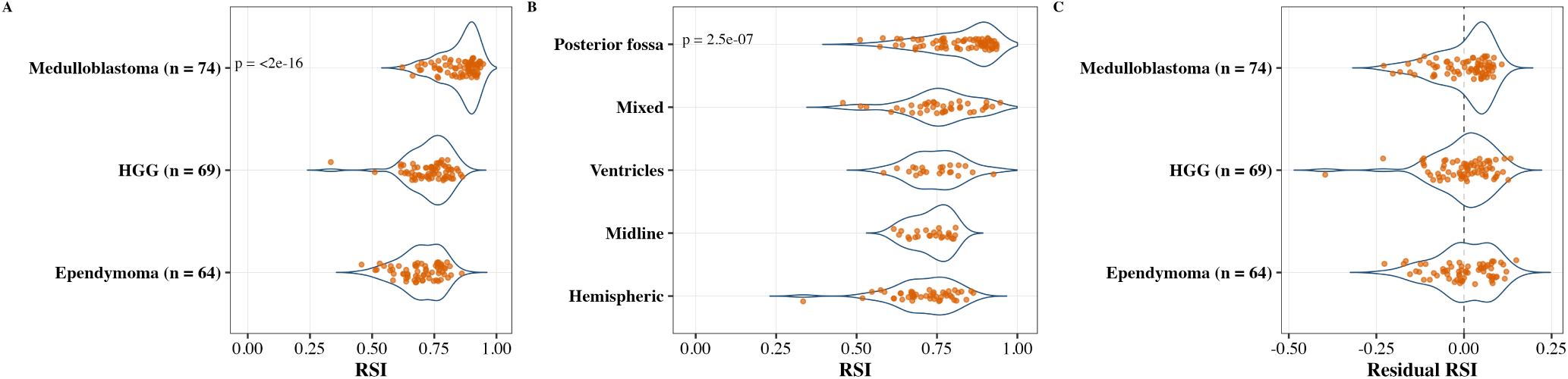
Relationship between clinical features and radiosensitivity index. **(A)** RSI distribution for each cancer type. **(B)** RSI distribution for each anatomic brain region. **(C)** Distribution of residuals for RSI following linear regression including both cancer type and anatomic brain region. p-value for all panels was computed using Kruskal-Wallis test. The heterogeneity in RSI residuals following subtype and brain region driven regression supports additional stratification.

### RSI Subtyping

The distribution of RSI was compared each tumor across clinically-relevant molecular subtypes. These analyses are presented in **Supplemental Figure 2**.

**Supplemental Figure 2.**
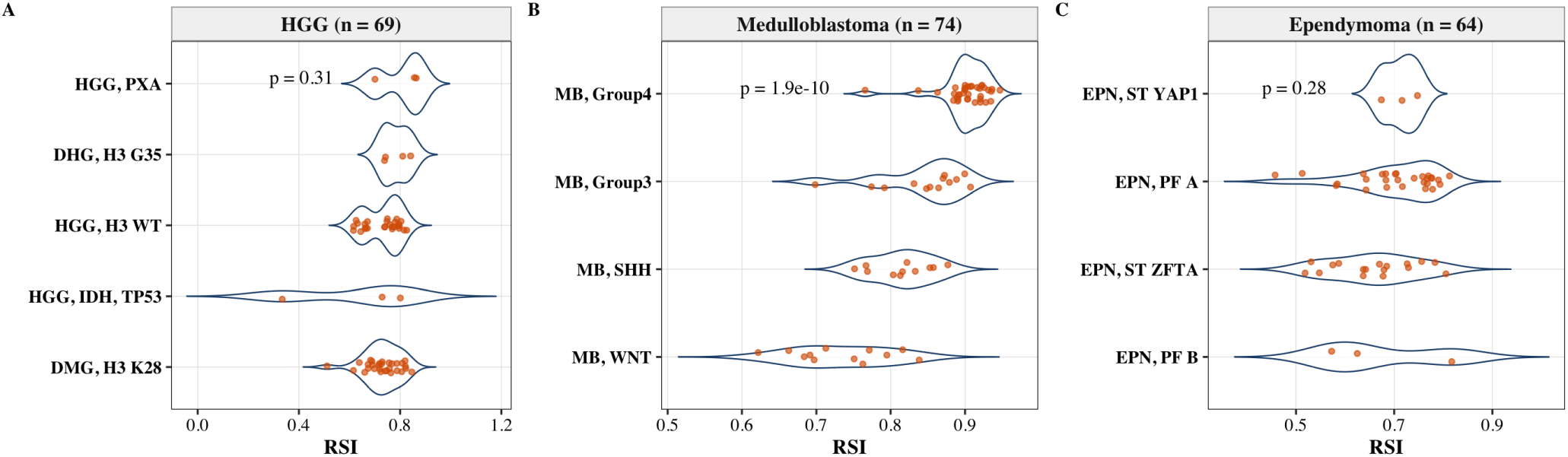
Variability in radiosensitivity across molecular subtypes of each tumor. The RSI distribution for **(A)** High-grade gliomas **(B)** Medulloblastomas and **(C)** Ependymomas are presented stratified by clinically-relevant molecular subtypes. p-value for all panels was computed using Kruskal-Wallis test.

### Physical RT dose and GARD distributions

Physical radiation therapy (RT) dose alone provides limited granularity for capturing differences in clinical outcomes at the individual patient level, particularly within the constraints of evidence-based practice where dosing is typically localized around similar values across trials. Incorporating a genomic framework through the genomic-adjusted radiation dose (GARD) enables quantification of substantial inter-patient variability in the biologic effect of RT, even when delivered doses span a narrow, uniform range. The distributions of physical RT dose and corresponding GARD values are presented in **Supplemental Figure 3**.

**Supplemental Figure 3.**
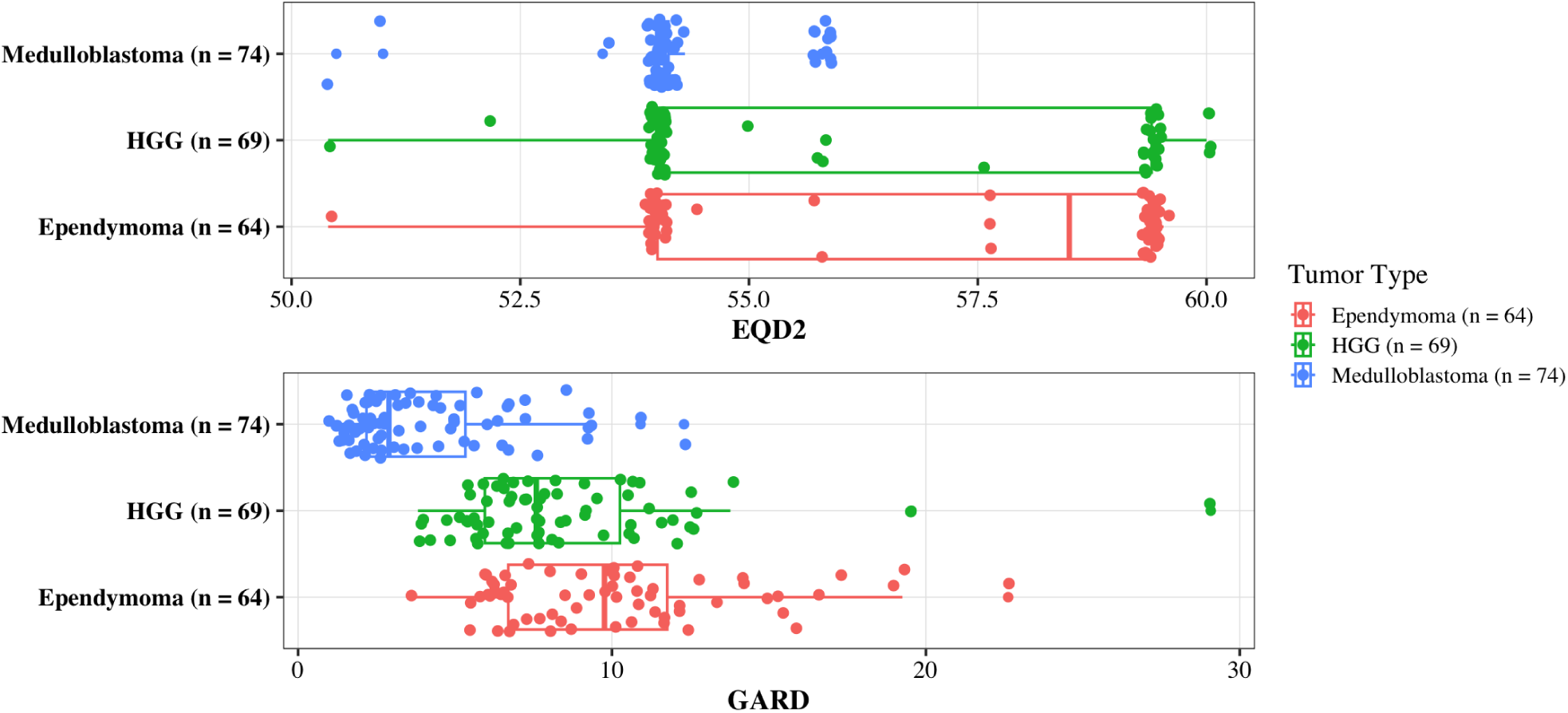
GARD and EQD2 distribution for each tumor type. EQD2 plot (top) measures total dose in Gy. GARD plot (bottom) measures biological effect in GARD units

### Relationship of clinical features with GARD

In **Supplemental Figure 4** we present the relationship between GARD and patient age and sex to assess potential associations.

**Supplemental Figure 4.**
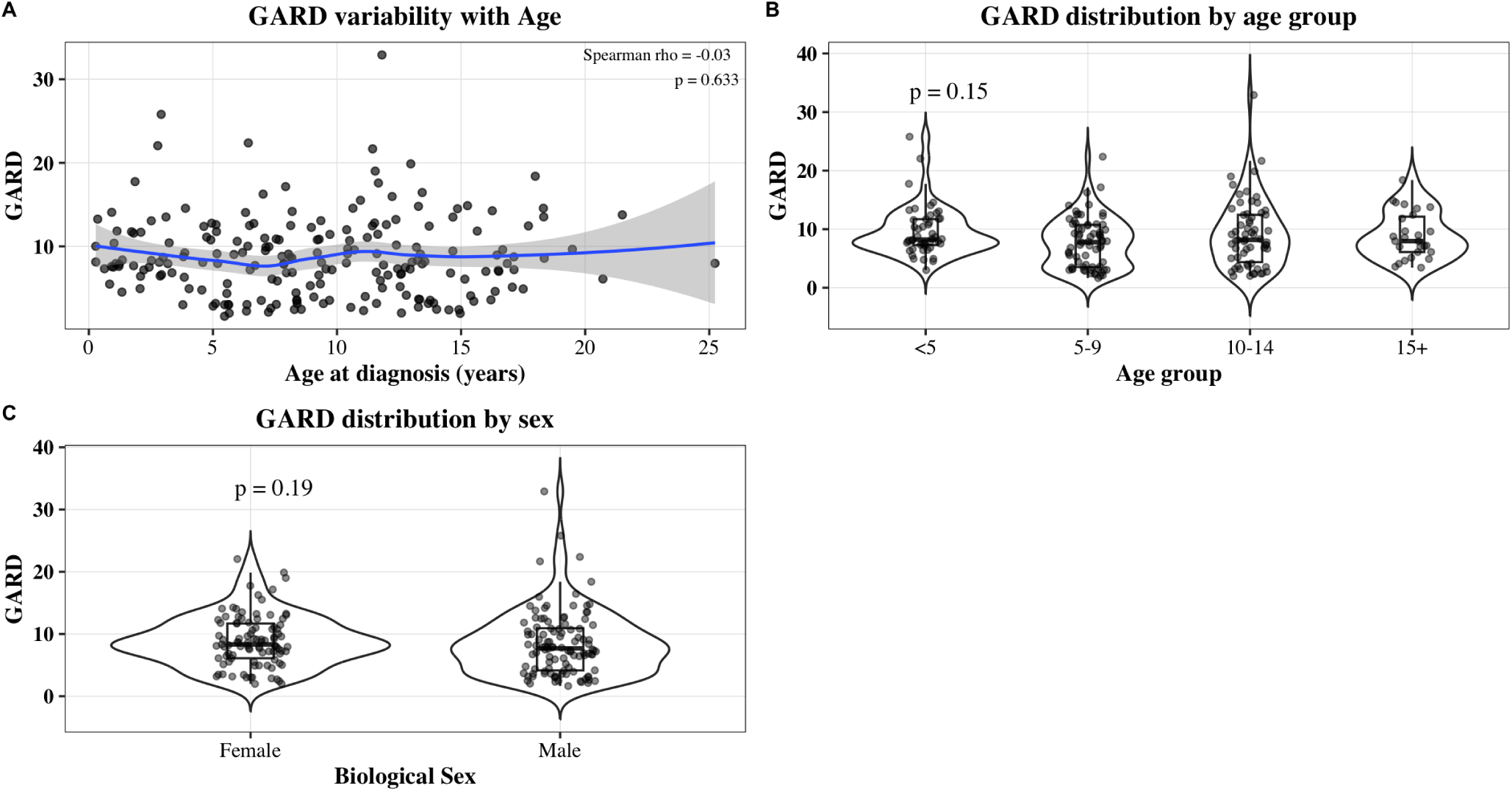
GARD and clinical feature distribution. **(A)** Correlation of GARD with age at diagnosis. Significance was evaluated using the Spearman correlation test. **(B)** Distribution of GARD across clinically meaningful age groups. Significance was evaluated using the Kruskal-Wallis test. **(C)** Distribution of GARD for biological sex. Significance was evaluated using the Wilcox test.

### Medulloblastoma GARD Distribution by Clinical Risk Group

Among patients with medulloblastoma who received radiotherapy, GARD distributions were compared between conventional clinical risk groups. High-risk and low-risk treatment groups generally received craniospinal irradiation doses of approximately 36 Gy and 23.4 Gy, respectively. Despite this difference in treatment intensity, GARD did not differ significantly between risk groups (*p* = 0.87), suggesting that clinical risk classification and prescribed CSI dose alone did not account for the observed variation in predicted biological effect. The distribution of molecular subtypes within each risk group is shown in **Supplemental Figure 5**.

**Supplemental Figure 5.**
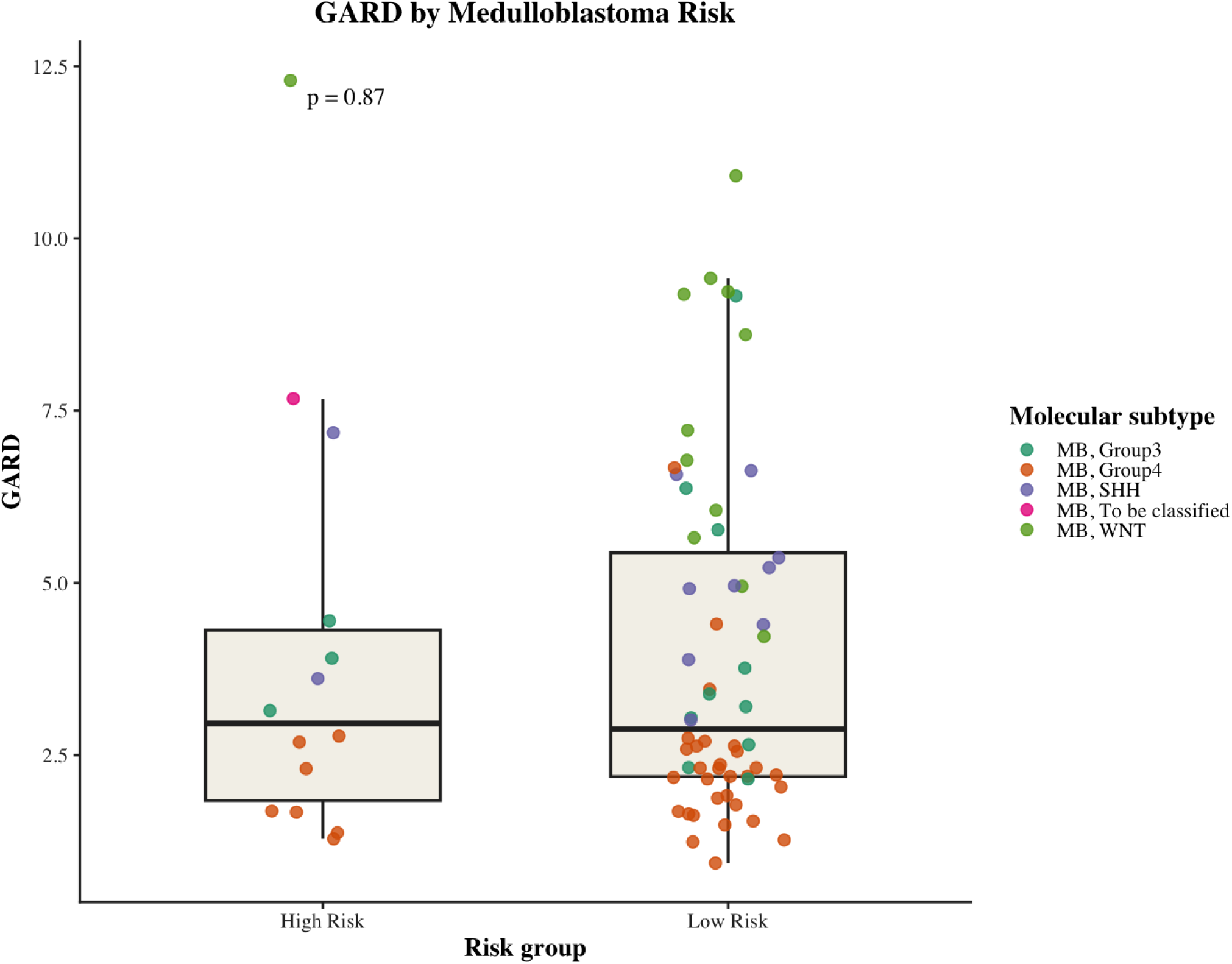
GARD distribution by medulloblastoma clinical risk group. GARD values are shown for patients with high-risk and low-risk medulloblastoma treated with radiotherapy. Box plots indicate the median and interquartile range, with whiskers extending to 1.5 times the interquartile range. Individual points represent patients and are colored according to molecular subtype. GARD distributions did not differ significantly between clinical risk groups (*p* = 0.87, two-sided Wilcoxon rank-sum test).

### Event-free Survival by Radiotherapy Status

In **Supplemental Figure 6** we present the Kaplan-Meier curves for recurrence for patients with and without RT, as applicable.

**Supplemental Figure 6.**
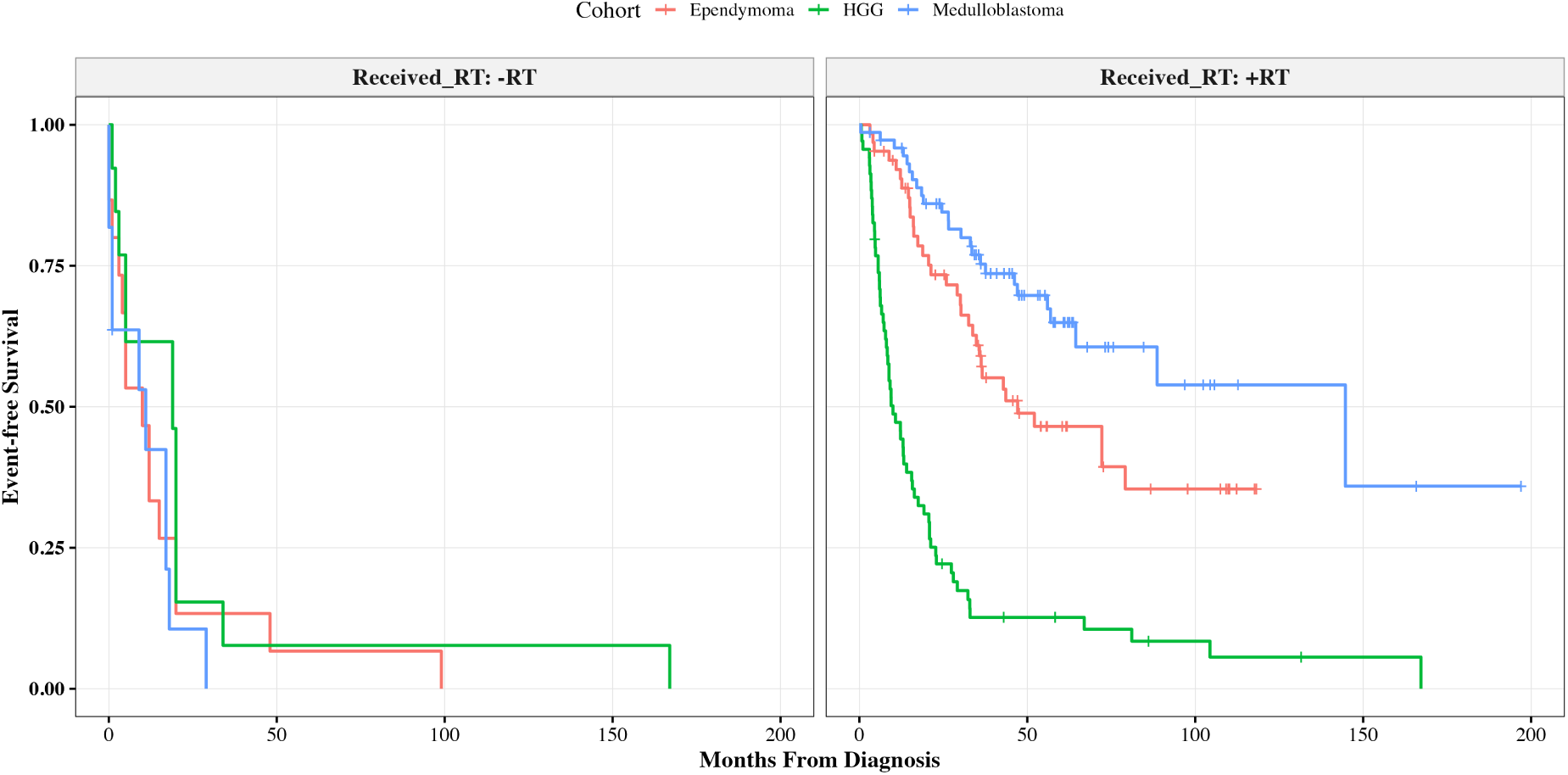
Kaplan-Meier curves for each cohort, both without (left) and with (right) RT with first recurrence as the outcome.

### Event-Free Survival by GARD

Event-free survival among patients receiving radiotherapy was evaluated according to GARD dichotomization. Kaplan–Meier curves for high-grade glioma, medulloblastoma, and ependymoma are presented in **Supplemental Figure 7**. Higher GARD was associated with improved event-free survival in medulloblastoma and ependymoma, whereas the difference did not reach statistical significance in high-grade glioma. Cutpoint was optimized for each histology using a minimum class proportion of 0.2.

**Supplemental Figure 7.**
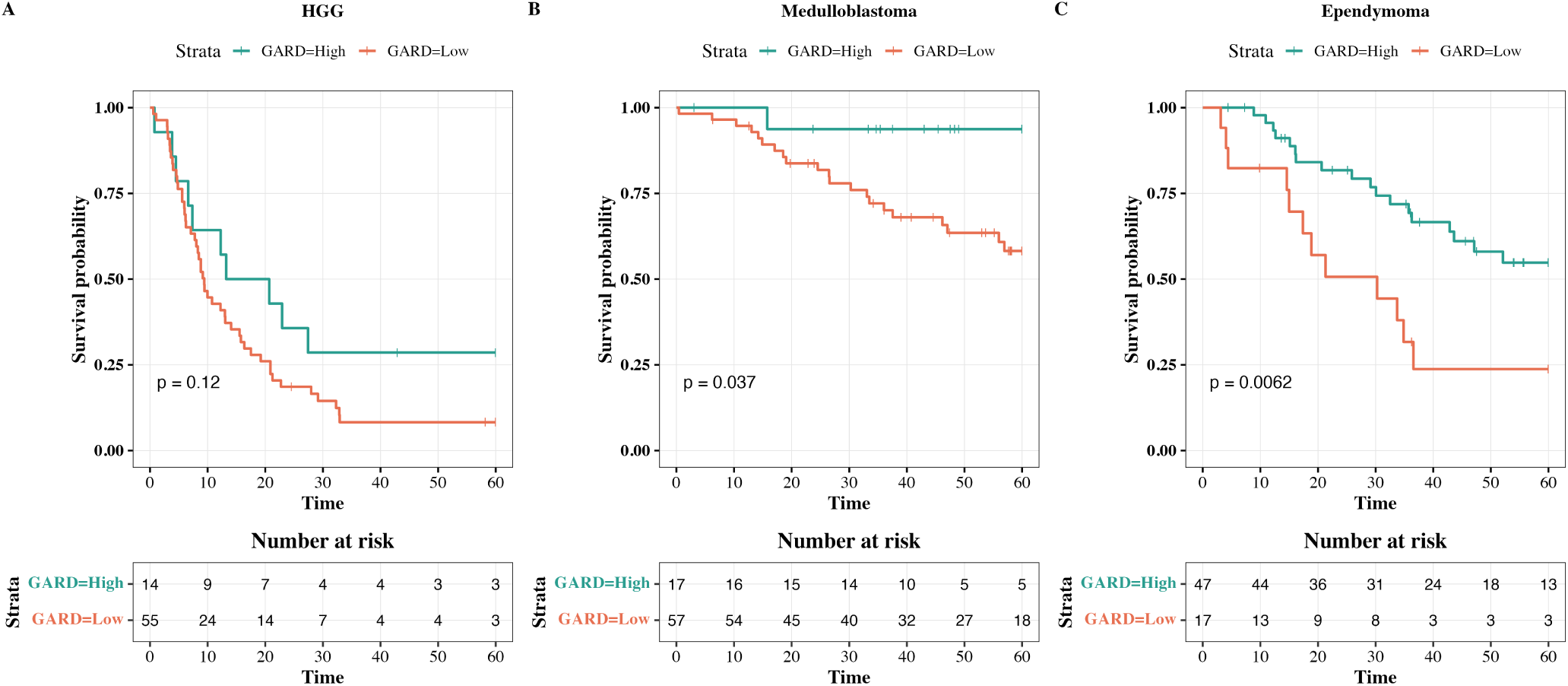
Event-free survival according to GARD group among patients receiving radiotherapy, stratified by tumor type. Kaplan–Meier curves compare patients with high versus low GARD within **(A)** high-grade glioma (HGG), **(B)** medulloblastoma, and **(C)** ependymoma. Tick marks indicate censored observations, and the corresponding numbers at risk are shown below each panel. Differences between survival curves were evaluated using two-sided log-rank tests.

### Overall Survival Cohort

In **Supplemental Figure 8** we present the Kaplan-Meier curves for overall survival for patients with and without RT, as applicable.

**Supplemental Figure 8.**
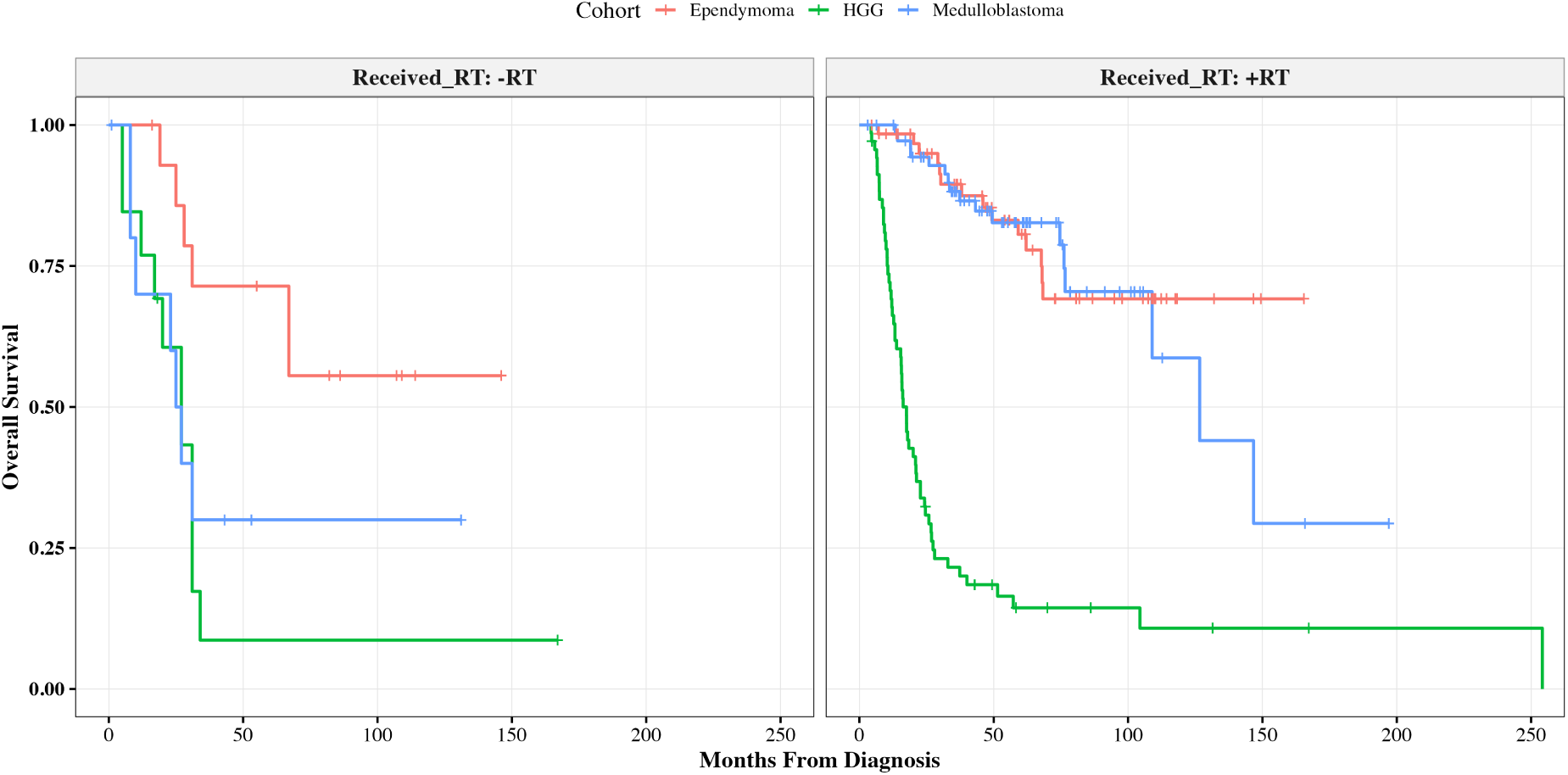
Kaplan-Meier curves for each cohort, both without (left) and with (right) RT with first overall survival as the outcome.

### Overall Survival by GARD

Overall survival among patients receiving radiotherapy was evaluated according to GARD dichotomization. Kaplan-Meier curves for high-grade glioma, medulloblastoma, and ependymoma are presented in **Supplemental Figure 9**. Higher GARD was associated with improved overall survival in ependymoma, whereas the difference did not reach statistical significance in high-grade glioma or medulloblastoma. Cutpoint was optimized for each histology using a minimum class proportion of 0.2.

**Supplemental Figure 9.**
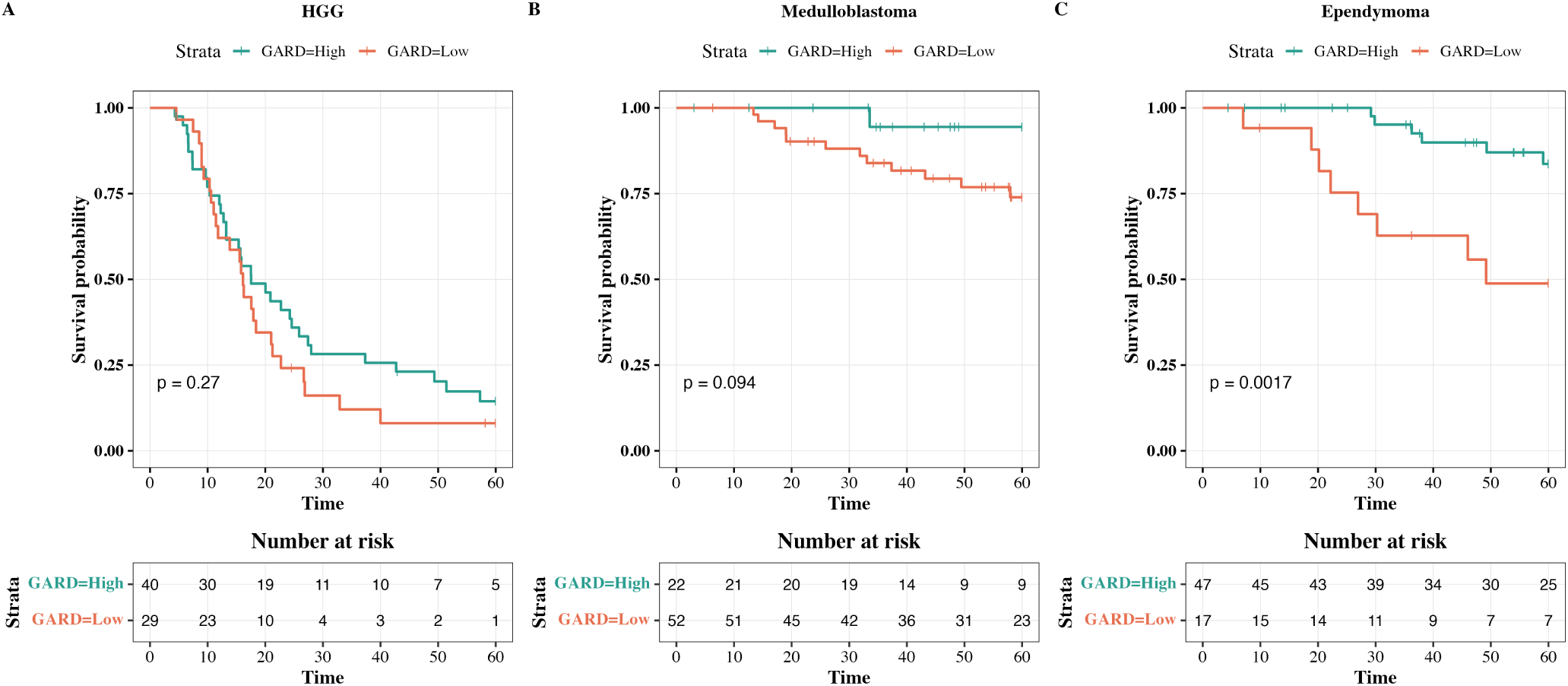
Overall survival according to GARD group among patients receiving radiotherapy, stratified by tumor type. Kaplan-Meier curves compare patients with high versus low GARD within **(A)** high-grade glioma (HGG), **(B)** medulloblastoma, and **(C)** ependymoma. Tick marks indicate censored observations, and the corresponding numbers at risk are shown below each panel. Differences between survival curves were evaluated using two-sided log-rank tests.

### Pooled Cox model results

In the following tables we describe the individual cox models for each of the forest plots presented in the manuscript. Separate tables are presented for GARD, sham-GARD, and Physical Dose.

#### GARD

GARD is significantly associated with both overall survival and recurrence in the pooled analysis (see **Supplemental Table 2**).

**Supplemental Table 2.** Hazard ratios for each disease site and outcome type with GARD as the covariate.

| Disease Site | Recurrence |  |  |  |  |  | Survival |  |  |  |  |  |
| --- | --- | --- | --- | --- | --- | --- | --- | --- | --- | --- | --- | --- |
|  | n | events | HR | lower | upper | p | n | events | HR | lower | upper | p |
| Ependymoma (n = 64) | 64 | 30 | 0.910 | 0.808 | 1.025 | 0.113 | 64 | 14 | 0.957 | 0.805 | 1.137 | 0.611 |
| HGG (n = 69) | 69 | 59 | 0.902 | 0.813 | 1.001 | **0.047** | 69 | 59 | 0.902 | 0.814 | 1.000 | **0.045** |
| Medulloblastoma (n = 74) | 74 | 22 | 0.846 | 0.675 | 1.060 | 0.137 | 74 | 13 | 0.807 | 0.594 | 1.097 | 0.163 |
| Pooled | 207 | 111 | 0.898 | 0.834 | 0.967 | **0.004** | 207 | 86 | 0.905 | 0.831 | 0.985 | **0.018** |

#### Physical RT Dose

Physical RT dose (**Supplemental Table 3**) has no statistically significant associations with any outcome.

**Supplemental Table 3.** Hazard ratios for each disease site and outcome type with EQD2 as the covariate.

| Disease Site | Recurrence |  |  |  |  |  | Survival |  |  |  |  |  |
| --- | --- | --- | --- | --- | --- | --- | --- | --- | --- | --- | --- | --- |
|  | n | events | HR | lower | upper | p | n | events | HR | lower | upper | p |
| Ependymoma (n = 64) | 64 | 30 | 0.906 | 0.777 | 1.056 | 0.197 | 64 | 14 | 1.188 | 0.929 | 1.519 | 0.161 |
| HGG (n = 69) | 69 | 59 | 1.023 | 0.916 | 1.143 | 0.677 | 69 | 59 | 1.013 | 0.908 | 1.131 | 0.811 |
| Medulloblastoma (n = 74) | 74 | 22 | 1.008 | 0.607 | 1.673 | 0.975 | 74 | 13 | 1.050 | 0.514 | 2.143 | 0.892 |
| Pooled | 207 | 111 | 0.981 | 0.897 | 1.073 | 0.676 | 207 | 86 | 1.042 | 0.946 | 1.149 | 0.395 |

#### Sham-GARD

Sham-GARD was not associated with recurrence or survival in the pooled cohort (**Supplemental Table^∼^??**{tab: supp*_s_hamgard*)*.Amongthedise speci f icanalyses, anisolatednominalassociationwithrecurrencewasobservedinthesmallmedulloblastomasubgroup*(*HR* = 1.92, 95%*CI*1.00 *−*3.69;p = 0.046)*, butnotwithoverallsurvival. This finding should be interpreted cautiously given the limited sample size*.

**Supplemental Table 4.** Hazard ratios for each disease site and outcome type with sham-GARD as the covariate.

| Disease Site | Recurrence |  |  |  |  |  | Survival |  |  |  |  |  |
| --- | --- | --- | --- | --- | --- | --- | --- | --- | --- | --- | --- | --- |
|  | n | events | HR | lower | upper | p | n | events | HR | lower | upper | p |
| Ependymoma (n = 15) | 15 | 14 | 0.877 | 0.706 | 1.090 | 0.228 | 15 | 6 | 1.049 | 0.767 | 1.437 | 0.759 |
| HGG (n = 13) | 13 | 12 | 0.739 | 0.472 | 1.157 | 0.177 | 13 | 12 | 0.823 | 0.528 | 1.283 | 0.380 |
| Medulloblastoma (n = 11) | 11 | 10 | 1.918 | 0.998 | 3.688 | **0.046** | 11 | 9 | 1.130 | 0.884 | 1.444 | 0.319 |
| Pooled | 39 | 36 | 0.973 | 0.831 | 1.140 | 0.733 | 39 | 27 | 1.042 | 0.875 | 1.242 | 0.638 |

### Sensitivity Analyses

To assess whether GARD is independently associated with survival beyond established disease features, we evaluated three Cox modeling approaches: an unadjusted model including GARD alone, a stratified model accounting for histology and anatomic brain region, and a multivariable model including these variables as covariates. Multivariable models demonstrate GARD is predictive for survival with histology and brain region included as covariates. (**Supplemental Table 4**, **Supplemental Figure 10**)

**Supplemental Table 5.** Association of GARD with recurrence and survival outcomes across modeling strategies.

| Model | Recurrence |  |  |  |  |  | Survival |  |  |  |  |  |
| --- | --- | --- | --- | --- | --- | --- | --- | --- | --- | --- | --- | --- |
|  | n | Events | HR | Lower | Upper | p | n | Events | HR | Lower | Upper | p |
| Unadjusted | 207 | 111 | 1.010 | 0.971 | 1.052 | 0.610 | 207 | 86 | 1.008 | 0.963 | 1.055 | 0.723 |
| Adjusted (Histology + Region) | 207 | 111 | 0.912 | 0.852 | 0.976 | **0.007** | 207 | 86 | 0.916 | 0.846 | 0.992 | **0.027** |
| Stratified | 207 | 111 | 0.898 | 0.834 | 0.967 | **0.004** | 207 | 86 | 0.905 | 0.831 | 0.985 | **0.018** |

**Supplemental Figure 10.**
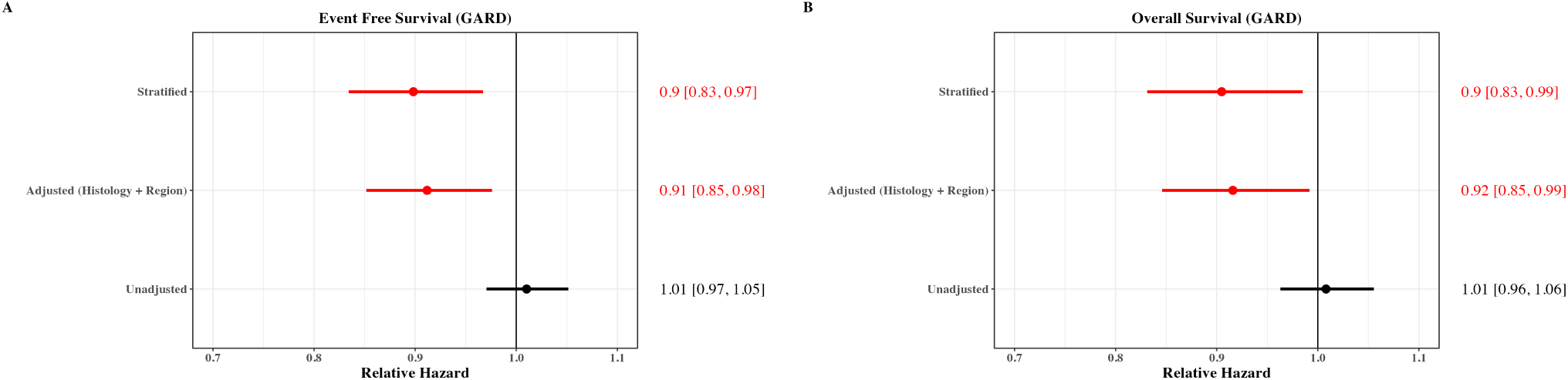
GARD is predictive for survival and recurrence across modeling strategies.

### Stratified Regression Results

#### Recurrence and Survival with and without RT

Stratified Cox proportional hazards models were constructed to evaluate the association between GARD and both event-free survival (EFS) and overall survival (OS), modeling GARD as a continuous linear variable in patients treated with and without radiation therapy. These analyses mirror the results shown in **Figure** 4, excluding the nomogram and instead using a linear modeling approach.

Among patients receiving radiation therapy, GARD was significantly associated with improved EFS (coefficient = *−*0.1073, *χ*^2^ = 8.41, p = 0.0037), whereas no such association was observed in patients who did not receive radiation (coefficient = *−*0.0270, *χ*^2^ = 0.1168, p = 0.7326; **Supplemental Figure** 11, left).

Similarly, GARD was significantly associated with OS in radiation-treated patients (coefficient = *−*0.0999, *χ*^2^ = 5.5483, p = 0.0185), but not in those who did not receive radiation (coefficient = 0.0412, *χ*^2^ = 0.2211, p = 0.6382; **Supplemental Figure 11**, right).

**Supplemental Figure 11.**
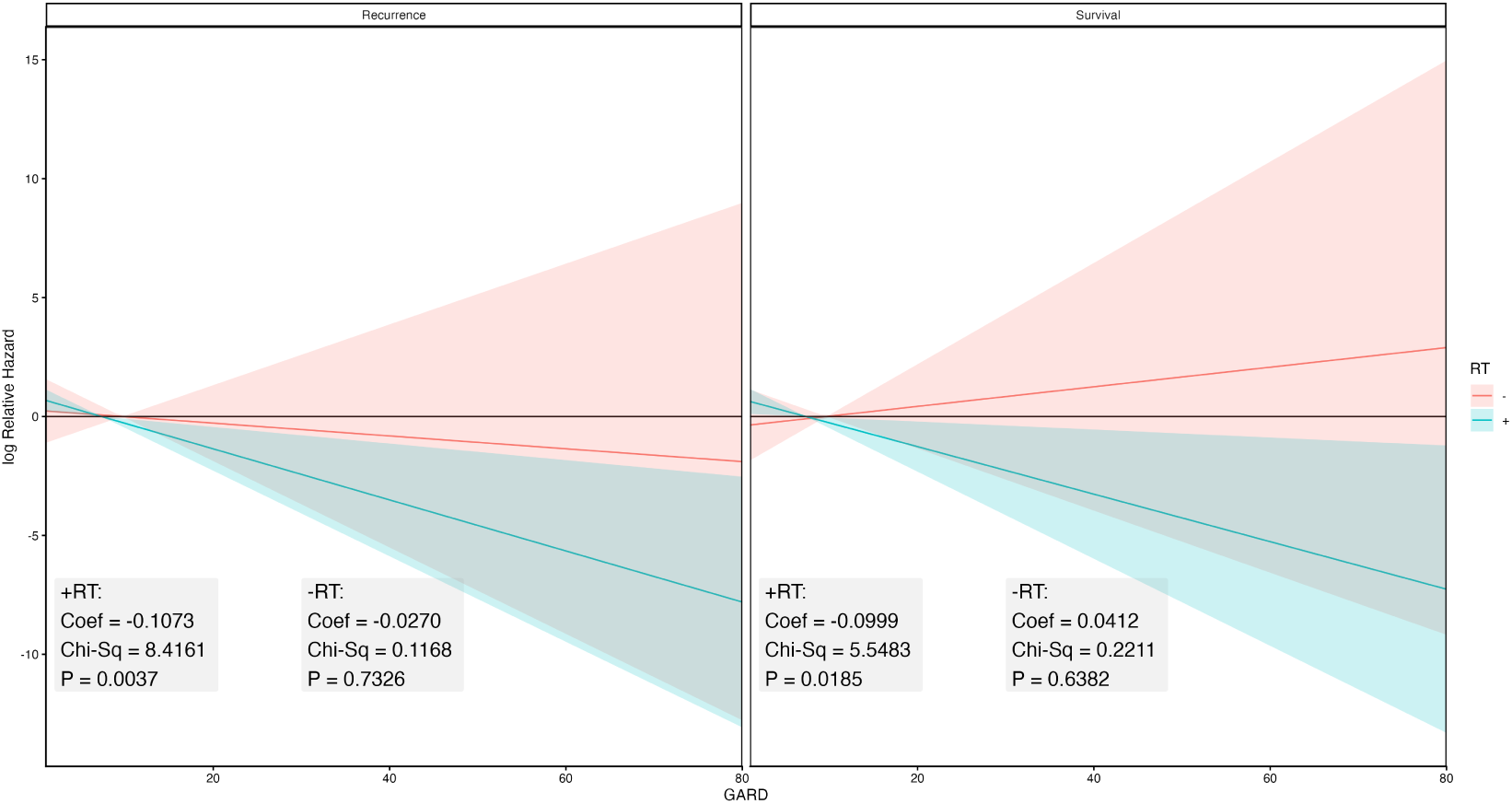
Stratified Cox regression analysis of HR for first recurrence (left) and overall survival (right) as function of GARD.

### Conflict of Interest

SAE and JTR hold patents and are co-inventors, co-founders and stock holders of Cvergenx, Inc. SAE is board member of Cvergenx, Inc. JGS holds patents and is a stock holder of Cvergenx, Inc.

## Notes

### Author Declarations

Source data is available at Children's Brain Tumor Network.

### Funding

- National Cancer Institute: U54-CA274513

